# Genetic liability, environmental adversity, and brain development predict adolescent polysubstance use initiation

**DOI:** 10.1101/2025.03.21.25324407

**Authors:** Dongmei Zhi, Brandon T. Sanzo, Daniel H. Jung, Judit Cabana-Domínguez, Noelia Fernàndez-Castillo, Bru Cormand, Jing Sui, Rongtao Jiang, 23andMe Research Team, Harrison T. Reeder, Sarah A. Thomas, A. Eden Evins, Scott E. Hadland, Joshua L. Roffman, Richard T. Liu, Jodi M. Gilman, Phil H. Lee

## Abstract

Polysubstance use (PSU) initiation often occurs during adolescence and is associated with increased risk of psychiatric comorbidity and substance use disorders (SUDs), yet factors associated with PSU initiation remain poorly understood. Using the Adolescent Brain Cognitive Development Study (N=9,174; 6-year follow-up), we examined associations of PSU initiation with shared genetic liability, multidimensional environmental exposures, and functional brain connectivity. We found that polygenic risk for shared genetic vulnerability across multiple SUDs, derived from independent adult GWAS, was associated with increased likelihood of PSU initiation. Environmental risk factors for PSU included peer victimization and adverse life events, particularly among adolescents with high genetic susceptibility, whereas parental monitoring and family cohesion were protective. Adolescents who later initiated PSU showed altered subcortical–cortical functional connectivity across development. These findings suggest that adolescent PSU initiation may mark a broader pattern of developmental vulnerability associated with shared genetic liability, environmental adversity, and altered brain network development.

## INTRODUCTION

Polysubstance use (PSU) initiation, defined as the initiation of multiple psychoactive substances within a one-year period^1^, is common during adolescence and is a major public health concern. More than 20% of U.S. adolescents report past-year PSU^2^, and early onset is associated with more severe and enduring consequences than single substance use (SSU), including higher rates of psychiatric comorbidity^3^ and increased risk for substance use disorders (SUDs) in adulthood^4,5^. Despite these consequences, most research has focused on individual substances rather than the broader pattern of PSU^6^. As a result, it remains unclear to what extent known risk factors for substance use specifically predispose certain youth to escalate from no substance use or SSU to PSU. Identifying early multimodal markers of vulnerability is critical for detecting adolescents at risk and informing targeted prevention during adolescence^7^.

Understanding PSU initiation requires examining multiple domains of risk that emerge before substance use begins. Genome-wide association studies (GWAS) have identified numerous loci associated with substance use and addiction vulnerability^8–10^ and have revealed substantial genetic overlap across SUDs^11,12^. Recent work has identified a latent “addiction risk factor” capturing shared genetic liability across alcohol, tobacco, cannabis and opioid use disorders, suggesting a common genetic vulnerability underlying multiple forms of substance use^11^. Critically, this general addiction polygenic risk score (PRS), independent of substance-specific genetic risk, has been associated with neuropsychiatric traits in adults^11,13^. However, whether and to what extent this shared genetic liability contributes specifically to the emergence of adolescent PSU remains unknown.

Environmental exposures also play a pivotal role in shaping the trajectory of substance use during youth^14–17^. Previous research has shown that adverse environments, such as traumatic events^16^ and peer substance use^17^, are risk factors for substance use in adolescents, whereas protective environments including high parental monitoring^15^, and strong family bonds^16^ may buffer against substance use initiation. However, few studies have assessed the effects of multidimensional (individual, family, school, and societal) environmental exposures on substance use in large adolescent cohorts, particularly with respect to PSU initiation. Notably, studies have shown gene-by-environment interactions in which environmental factors may buffer or amplify risks of substance use among individuals with high genetic susceptibility^18^. Yet most prior studies have examined environmental exposures individually or within limited domains, and research investigating how multiple environmental exposures interact with genetic susceptibility to influence PSU initiation in adolescence remains limited.

Neurodevelopmental processes may also contribute to vulnerability for substance use. Altered functional connectivity (FC) within cortico-striatal-thalamic circuits has been observed in individuals with SUDs, implicating networks involved in reward processing and executive control^19–21^. Studies of adults who engage in concurrent substance use suggest that PSU may be associated with distinct disruptions in brain network organization compared with SSU^20^. For example, adults engaging in concurrent use of alcohol and cannabis exhibit hyperconnectivity in salience and temporal networks compared to those using alcohol alone^20^, suggesting that PSU specifically may be linked to distinct disruptions in brain networks. However, most of this work has been conducted in cross-sectional samples of adults, after substance use patterns are already established. As a result, it remains unclear whether these neural differences reflect pre-existing vulnerabilities that predispose individuals to initiate PSU or neuroadaptations that emerge following substance exposure. Longitudinal studies beginning prior to substance use initiation are therefore critical for determining whether alterations in brain network development precede or follow the onset of polysubstance use during adolescence, directly addressing prior calls for developmentally informed longitudinal research in substance use.

Here, we address these gaps using data from the Adolescent Brain Cognitive Development (ABCD) Study, a large population-based longitudinal cohort with repeated assessments of substance use, genetic variation, environmental exposures and functional network connectivity. Specifically, we tested four related hypotheses. First, we examined whether shared genetic liability for adult SUDs associated with PSU initiation in adolescents. Second, we tested whether environmental adversity increases risk for PSU initiation whereas protective environmental factors reduce that risk. Third, we evaluated whether environmental exposures moderate the association between genetic liability and PSU initiation. Fourth, we examined whether adolescents who initiate PSU show differences in the developmental trajectory of functional network connectivity before polysubstance exposure. Together, this study aimed to identify early markers of vulnerability and clarify the extent to which genetic risk, environmental experiences, and brain network development jointly contribute to PSU initiation during adolescence.

## RESULTS

The sample consisted of 9,174 participants (4,396 females [47.9%]; mean age at baseline 9.95 [standard deviation 0.62] years; **Table 1**) with available lifetime substance use outcomes from the ABCD Study (**Methods and Extended Data Fig. 1**). Analysis included data from baseline to the first tranche of 6-year follow-up, with the remaining 6-year data available in subsequent releases^22^ (**Supplementary Table 1**). All analyses adjusted for the same biological, design-related, and socioeconomic factors (**Methods**).

**Table 1.**
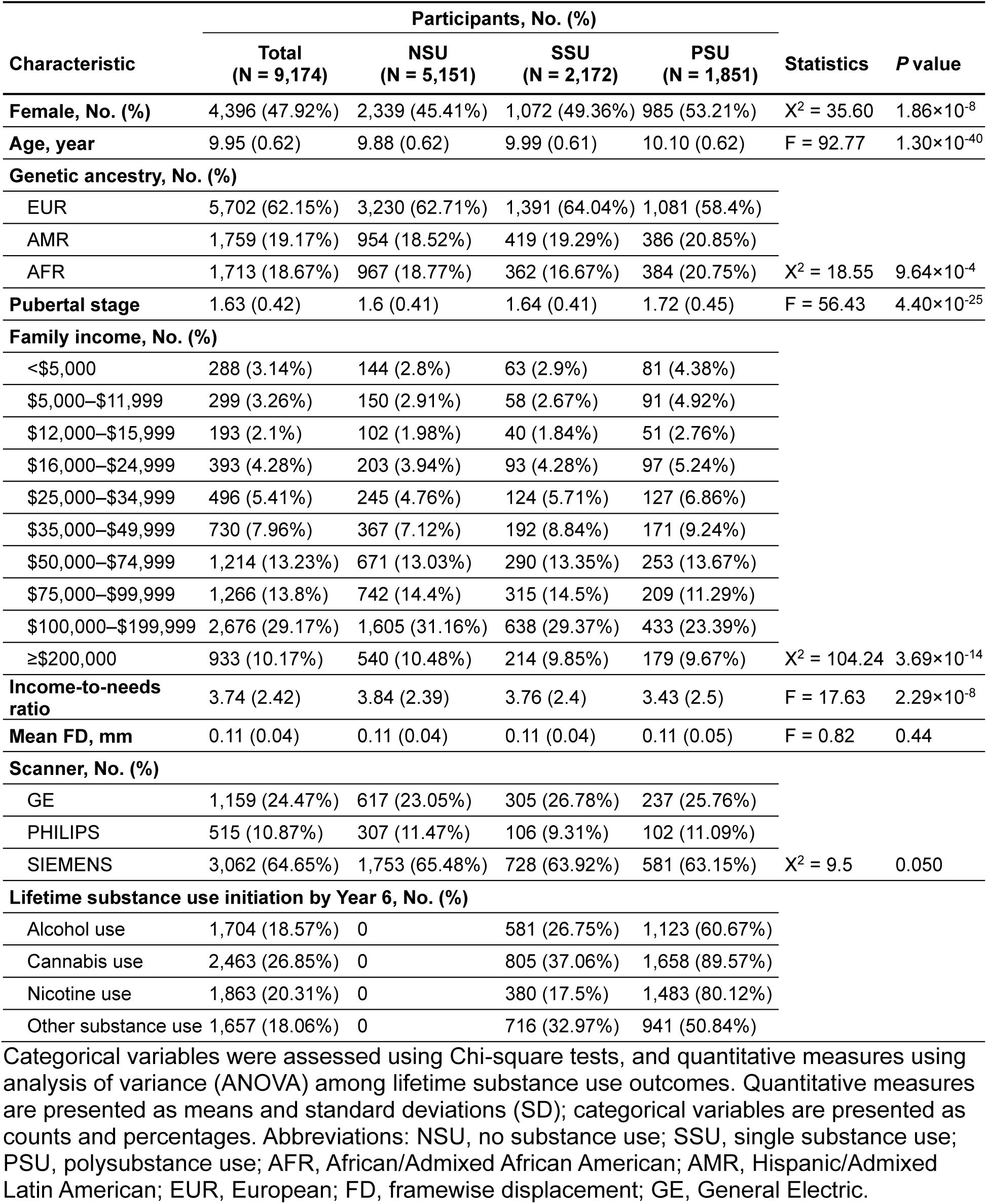
Baseline characteristics for participants stratified by lifetime substance use outcomes.

### Adolescents PSU Initiation and Risk for Psychopathology

Substance use was measured every 6 months^23^. Of the 9,174 participants, only 1.5% reported any substance use at baseline (age 9-10), but 2,172 (23.7%) reported lifetime SSU initiation and 1,851 (20.2%) reported PSU initiation, defined as concurrent use of at least two types within a one-year period^1^, by 6-year follow-up (**Fig. 1a; Extended Data Fig. 2**). Among the four substances examined, cannabis (2,463 [26.9%]) and nicotine (1,863 [20.3%]) were the most frequently reported and were also the most commonly co-used within the PSU group (**Figs. 1b-c**). Most transitions from NSU to PSU initiation occurred between the 4- and 6-year follow-up, with relatively short transition times from SSU to PSU (**Fig. 1d**). Compared with adolescents with SSU or NSU, adolescents who developed PSU showed persistently higher Child Behavior Checklist (CBCL) total problems, excluding three substance-related items, across all time points, adjusting for the listed covariates (β=0.059 to 0.185, T value=3.63 to 12.63, P_FDR_<0.05; **Fig. 1c**; **Methods; Supplementary Tables 4-5**).

**Fig. 1.**
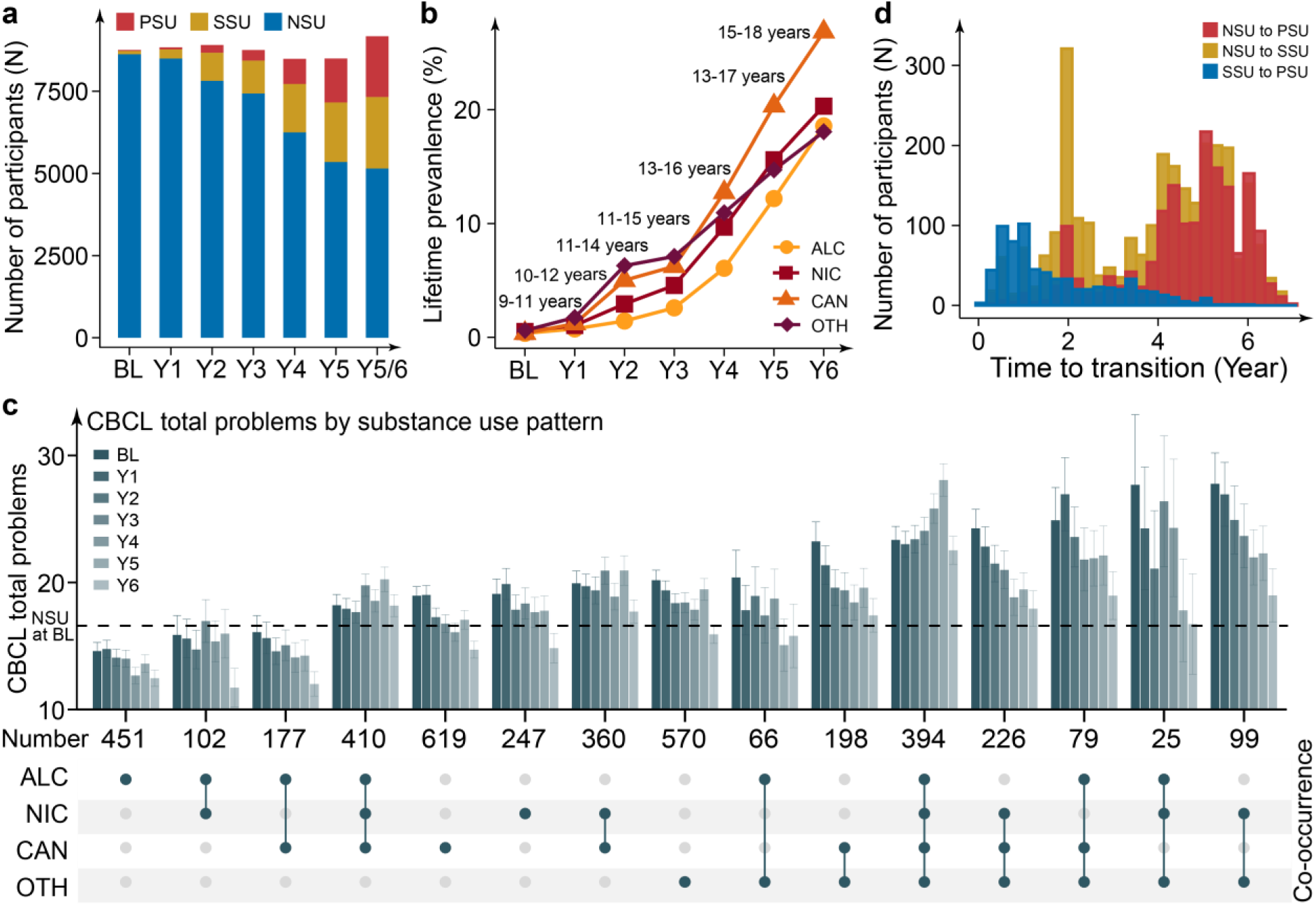
Lifetime substance use prevalence and their associations with psychopathology. (**a**) Lifetime prevalence of no substance use (NSU), single substance use (SSU), and polysubstance use (PSU) from baseline to 6-year follow-up. Given that 6-year follow-up data were available for roughly half of the cohort with the remaining 6-year data available in subsequent releases^22^, the most recent assessment at either 66-month or 5-year follow-up was used for participants without 6-year data, referred to as Y5/6 (**Extended Data** Fig. 1). (**b**) Lifetime prevalence of alcohol (ALC), nicotine (NIC), cannabis (CAN), and other substance use (OTH) from baseline to 6-year follow-up. The minimum and maximum ages of participants at each follow-up wave are provided in the text. (**c**) Mean and standard errors for CBCL total problems from baseline to 6-year follow-up across distinct substance use patterns. The x-axis shows distinct substance use patterns along with sample sizes. The dot matrix below indicates co-occurrence among four substances. The dashed line represents the mean baseline CBCL total problems for NSU group. (**d**) Transition time from NSU to PSU initiation—including individuals who transitioned through SSU, from NSU to SSU, and from SSU to PSU initiation. Most transitions from NSU to PSU initiation occurred between 4- and 6-year follow-up from baseline, with relatively short transition times from SSU to PSU initiation. BL, baseline; Y1: 1-year follow-up; Y2: 2-year follow-up; Y3: 3-year follow-up; Y4: 4-year follow-up; Y5: 5-year follow-up; Y6: 6-year follow-up.

### Associations of PRSs for SUDs with PSU

We first examined associations of one shared and five disorder-specific PRSs for SUDs with PSU (**Fig. 2a; Methods**; **Supplementary Tables 6-7**). In adjusted models, only PRS_SUD_, indexing shared genetic liability across five SUDs, was significantly associated with lifetime PSU compared to NSU in participants of European ancestry (EUR; OR=1.41, 95% CI=1.28–1.55, P_FDR_=2.03×10^−11^, Pseudo R^2^ = 2.35%) and Hispanic/Latin American ancestry (AMR; OR=1.40; 95% CI, 1.20–1.64; P_FDR_=2.33×10^−4^; Pseudo R^2^ = 2.70%; **Fig. 2b-c; Supplementary Table 8**). PRS_SUD_ also predicted shorter time to transition from NSU to PSU initiation in participants of EUR (HR=1.31, 95% CI=1.21–1.41, P_FDR_=1.23×10^−10^) and AMR (HR=1.25, 95% CI=1.10–1.43, P_FDR_=6.16×10^−3^; **Fig. 2d; Supplementary Table 9**). No PRSs were significantly associated with SSU compared with NSU (**Supplementary Table 10**).

**Fig. 2.**
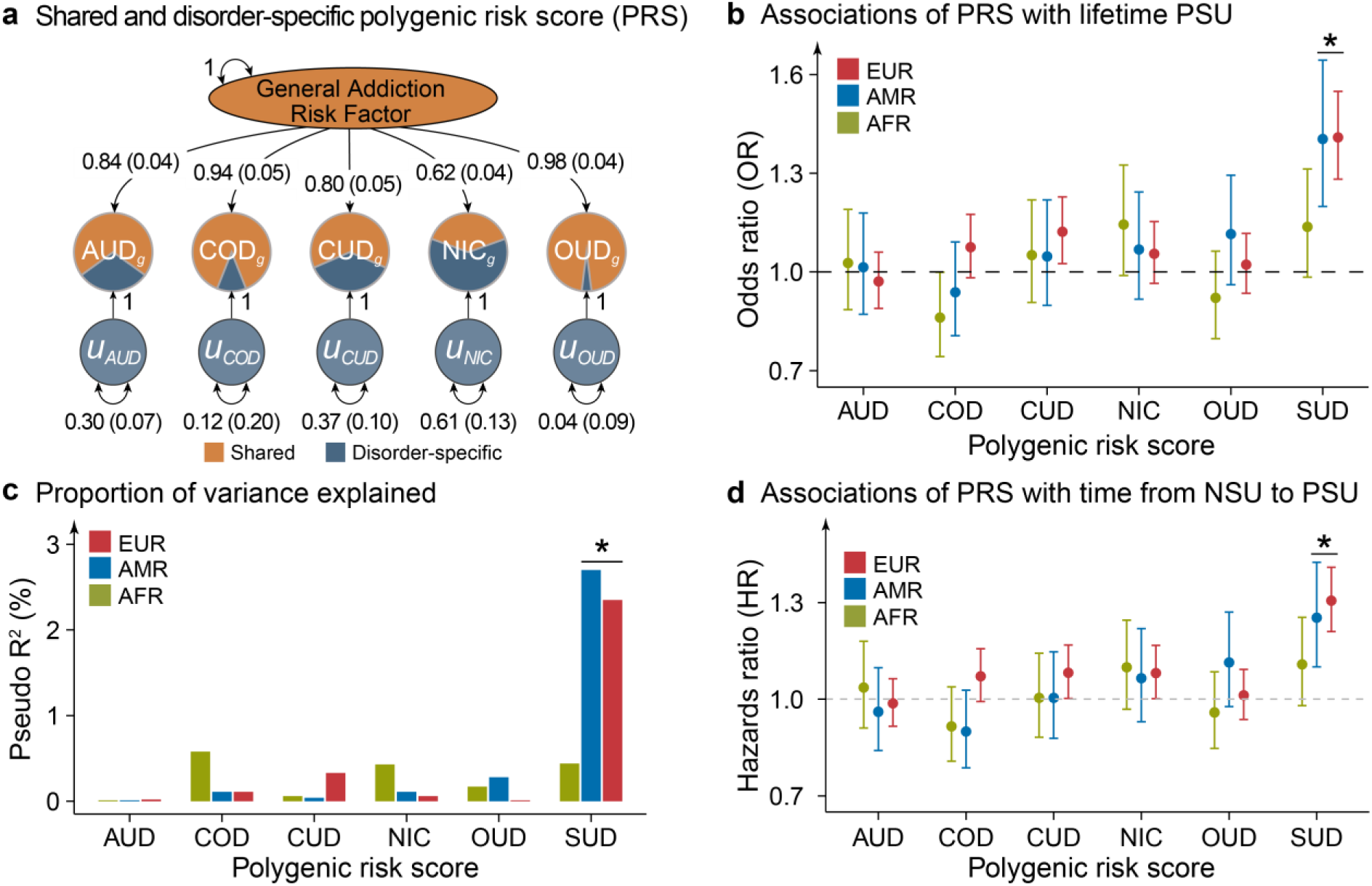
General addiction risk factor derived from genomic structural equation modeling (GenomicSEM). (**a**) The latent “General Addiction Risk Factor”, capturing the common genetic liability across five substance use disorder (SUDs), was estimated using GenomicSEM based on genome-wide association summary statistics for alcohol use disorder (AUD), cocaine use disorder (COD), cannabis use disorder (CUD), nicotine dependence (NIC), and opioid use disorder (OUD). Standardized estimates from the GenomicSEM model are shown, with standard errors in parentheses. Estimates are standardized relative to SNP-based heritability, such that each disorder’s genetic variance is decomposed into the variance explained by the general addiction risk factor (the squared factor loadings; single-headed arrows from the general addiction risk factor to each disorder) and residual variance (values on the double-headed arrows for the residual terms, labelled as *u*). SUDs are depicted as pie charts, where the proportion of variance explained by the general addiction factor is shown in orange, and the residual (disorder-specific) genetic variance is shown in blue. (**b**) Associations between six polygenic risk scores (PRSs) for addiction risk and lifetime PSU by 6-year follow-up within each genetic ancestry group, shown as adjusted odds ratios (ORs) with 95% confidence intervals (CIs), controlling for age, sex, pubertal stage, income-to-needs ratio, and random effects for family ID, and sites. (**c**) Pseudo R^2^ values explained by six PRSs for lifetime PSU within each genetic ancestry group. (**d**) Adjusted hazards ratio (HR) with 95% CIs for associations between six PRSs and time to transition from no substance use (NSU) to PSU initiation within each genetic ancestry group. *, P_FDR_ < 0.05, significant after false discovery rate (FDR) correction for 18 multiple comparisons (three genetic ancestry × six PRSs). Abbreviations: SUD, substance use disorder; EUR, European; AFR, African/Admixed African American; AMR, Hispanic/Admixed Latin American.

### Associations of Environment and PRS-by-Environment Interactions with PSU

To identify key environmental risk and protective exposures for PSU, we examined associations of 88 exposures at the initial assessment, after excluding highly colinear exposures (variance inflation factors >5), with substance use outcomes (**Methods; Extended Data Fig. 3**). Of these, 63 (72%) were significantly associated with lifetime PSU compared to NSU after adjusting for the listed covariates (risk factors: OR=1.08–1.52; protective factors: OR=0.58–0.91, P_FDR_<0.05; **Fig. 3a; Supplementary Table 11**). Specifically, the strongest risk factors for PSU included youth-reported adverse life events (OR=1.52, 95% CI=1.41–1.63, P_FDR_=1.43×10^−27^), peer reputational victimization (OR=1.50, 95% CI=1.40–1.61, P_FDR_=1.25×10^−28^), and R-rated screen use (OR=1.43, 95% CI=1.34–1.54, P_FDR_=1.47×10^−22^). In contrast, parental monitoring (OR=0.58, 95% CI=0.54–0.63, P_FDR_=3.83×10^−35^), youth-reported family cohesion (OR=0.70, 95% CI=0.65–0.75, P_FDR_=1.39×10^−22^), and longer sleep duration (OR=0.74, 95% CI=0.69–0.80, P_FDR_=1.00×10^−14^) were the strongest protective factors (**Fig. 3b**).

**Fig. 3.**
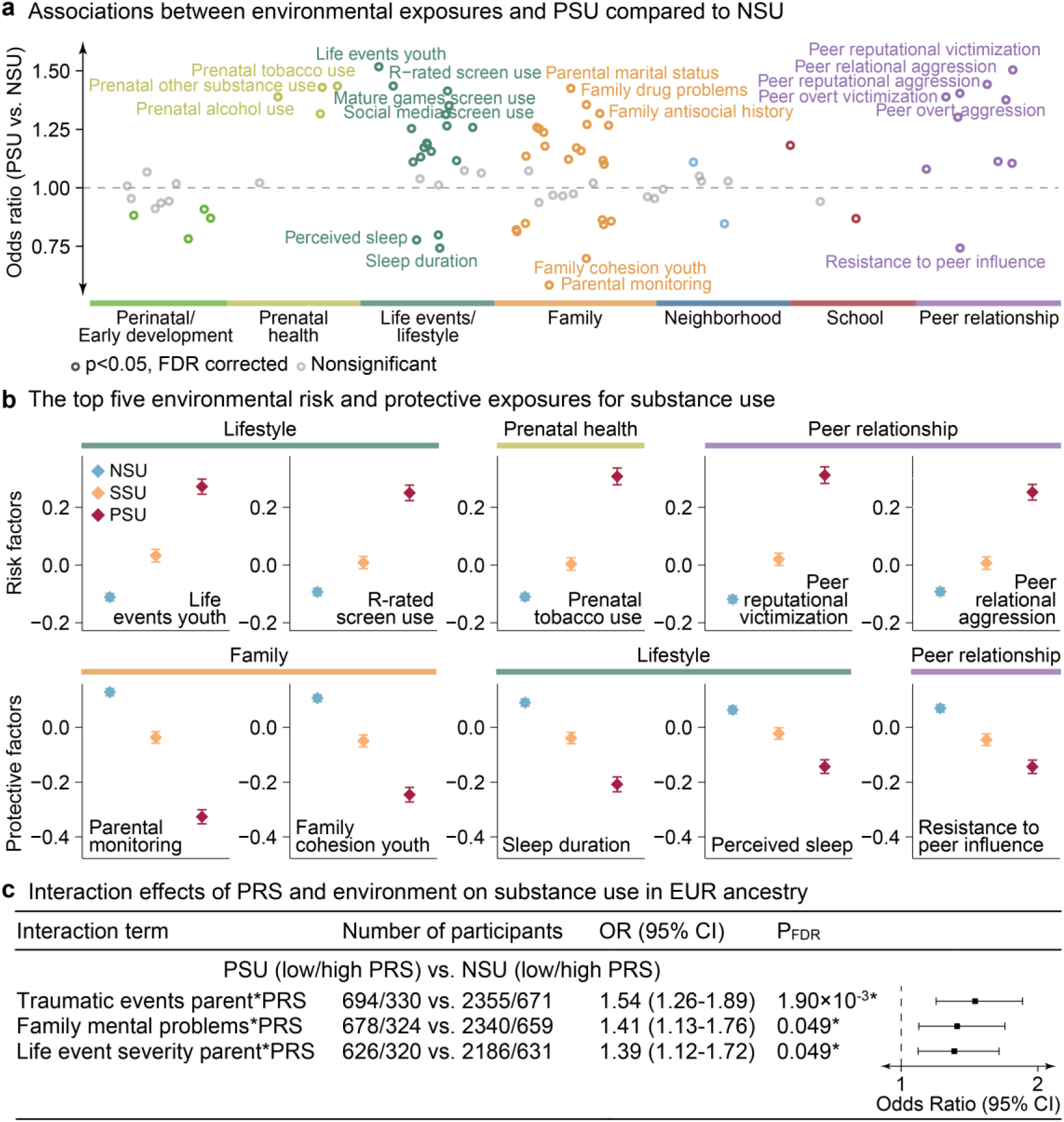
Associations between environmental exposures and polysubstance use (PSU). (**a**) Among 88 environmental exposures, 63 (72%) were significantly associated with PSU versus no substance use (NSU) after adjusting for age, sex, genetic ancestry, pubertal stage, income-to-needs ratio, and enrollment sites (P_FDR_<0.05, significant after false discovery rate (FDR) correction for 88 multiple comparisons). The fifteen strongest risk and the five most protective exposures are highlighted. Exposures without significant associations are shown in gray. (**b**) Mean and standard deviation values of the top five risk and top five protective environmental exposures showing the strongest associations with PSU versus NSU. All these exposures were significantly associated with both PSU versus NSU, and PSU versus single substance use (SSU). The Y-axis represents mean z-score for each exposure, while the X-axis denotes the three substance use groups: NSU, SSU, and PSU. Adolescents with PSU initiation were exposed to lower levels of protective exposures but higher levels of risk exposures than those with NSU, and SSU initiation. (**c**) Interaction effects of environmental exposures and genetic risk groups on PSU versus NSU in adolescents of European (EUR) ancestry, with high-genetic-risk group defined as the top 25% PRS_SUD_ and low-genetic-risk group as the bottom 75% PRS_SUD_^24,25^. Results for PSU versus SSU, and for Hispanic/Admixed Latin American (AMR) and African/Admixed African American (AFR) are presented in **Supplementary Tables 17–18**. Odds ratios (ORs) with 95% confidence intervals (CIs) are shown. The number of participants is presented for the PSU and NSU groups, stratified by low and high genetic risk. *, P_FDR_ < 0.05.

When grouped into environmental domains (**Methods**), peer relationships, life events or lifestyle, and prenatal exposures showed the largest proportions of significant associations, with most variables within these domains significantly associated with PSU. Notably, all environmental factors significantly associated with PSU versus SSU were also significant for PSU versus NSU, with the SSU group showing intermediate exposure levels between NSU and PSU (**Fig. 3b**). Associations of environment with lifetime SSU versus NSU, PSU versus SSU, and transition time from NSU to PSU (or SSU) initiation, and from SSU to PSU initiation showed similar findings (**Extended Data Figs. 4–5**, **Supplementary Tables 12–16**).

We next examined whether PRS_SUD_ modified associations between environmental exposures and PSU by stratifying participants into a high- (top 25% PRS_SUD_) and low-genetic-risk (bottom 75% PRS_SUD_) groups based on PRS_SUD_ within each genetic ancestry^24,25^. In adolescents of EUR, associations of traumatic events, family mental problems, and parent-reported life event severity with PSU versus NSU were stronger in the higher-genetic-risk group than in the low-risk group (P_FDR_ < 0.05; **Fig. 3c; Supplementary Table 17**). No significant interactions were observed for PSU versus SSU in adolescents of EUR, AMR, and AFR (**Supplementary Table 18**).

### Altered Subcortical-Cortical FC Associated with PSU

Participants underwent biennial magnetic resonance imaging scans (baseline and 2-, 4-, and 6-year follow-up)^26^. To characterize alterations in functional network connectivity over time among adolescents with lifetime PSU initiation, we estimated Fisher Z-transformed averaged FC for each participant within and between 13 Gordon cortical networks, and between 13 cortical networks and 19 subcortical regions^26^, after applying rigorous quality control and mean FD<0.2 mm (**Methods, Extended Data Fig. 1**). After covariates adjustment, among 247 subcortical-cortical FCs, the number of connectivity measures that significantly differed by lifetime PSU versus NSU increased with age across longitudinal scans (0 at baseline, 13 at 2-year, 121 at 4-year, and 82 at 6-year follow-ups (but note lower available sample size); increased FCs: OR=1.11–1.29; decreased FCs: OR=0.77 to 0.90, P_FDR_<0.05; **Fig. 4a**; **Supplementary Table 19**), indicating progressively greater network alterations in adolescents with PSU. Specifically, the most pronounced increased FCs for PSU versus NSU were observed between subcortical regions and transmodal association networks, particularly the fronto-parietal (FRP) and cingulo-parietal (CIP) networks. The strongest effects were observed for thalamic connections with these networks at the 4-year follow-up, including bilateral thalamus–CIP (right: OR=1.29, 95% CI=1.17–1.42, P_FDR_=2.04×10^−5^; left: OR=1.24, 95% CI=1.13–1.36, P_FDR_=1.99×10^−4^), and bilateral thalamus–FRP (right: OR=1.23, 95% CI=1.12–1.35, P_FDR_=3.18×10^−4^; left: OR=1.24, 95% CI=1.12–1.36, P_FDR_=2.06×10^−4^). In contrast, decreased FCs were primarily observed between subcortical regions and primary sensory and motor networks, including auditory (AUD), sensorimotor hand (SMH), and sensorimotor mouth (SMM) networks, as well as ventral attention (VEA) network. Among subcortical regions, the thalamus, caudate, and cerebellum were most consistently implicated in PSU, with 11 of 13 left caudate–cortical and 9 of 13 right thalamus–cortical connections significantly associated with PSU.

**Fig. 4.**
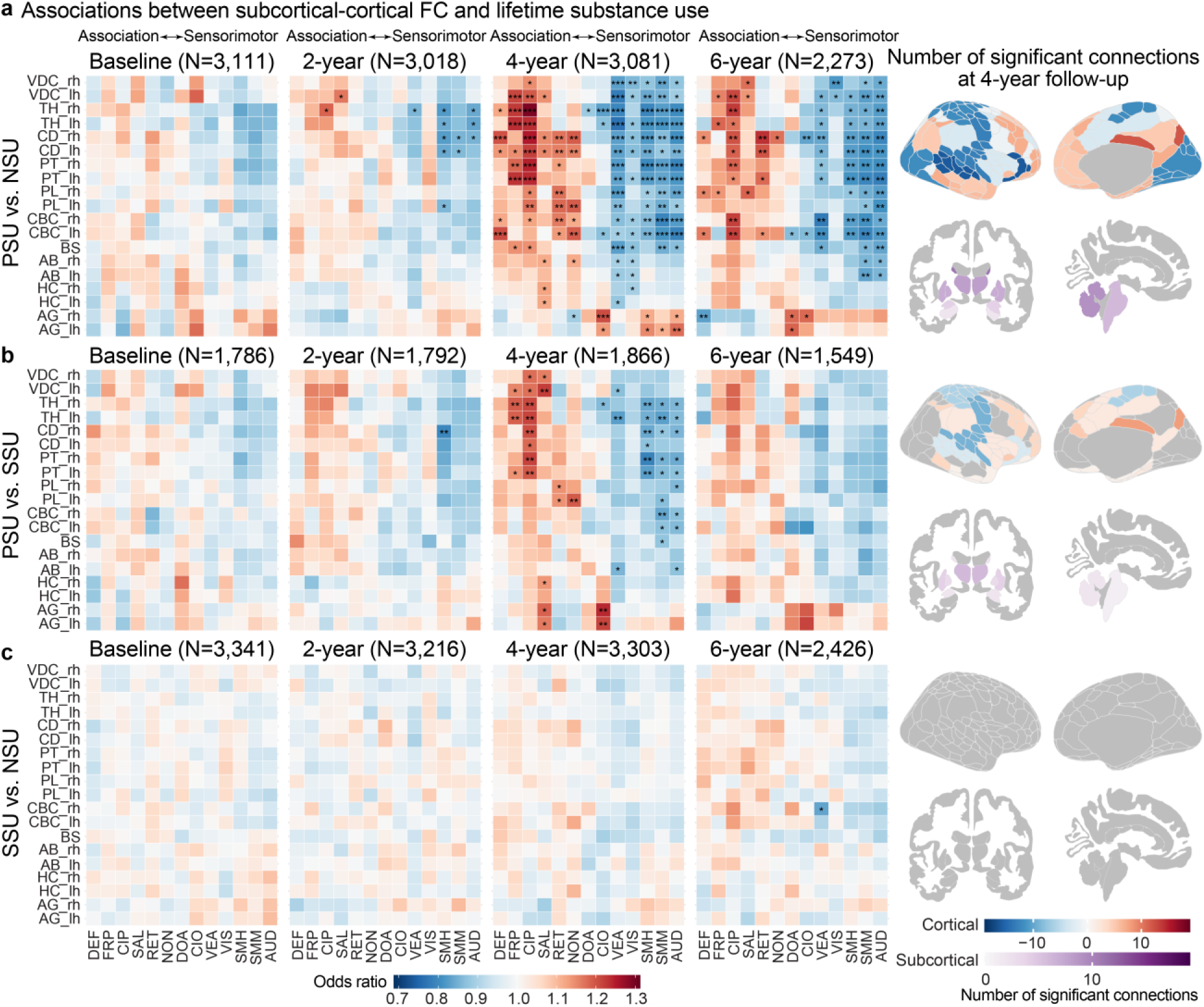
Associations between subcortical-cortical functional connectivity (FC) and lifetime substance use across time points. (**A**) Compared with no substance use (NSU), progressively more pronounced FC alterations were observed over time in lifetime polysubstance use (PSU) initiation by 6-year follow-up, with 0 significant associations at baseline, 13 at 2-year, 121 at 4-year, and 82 at 6-year follow-up, after adjusted for the listed covariates. Increased FCs were mainly observed between subcortical regions and transmodal association cortical networks, particularly the thalamus–cingulo-parietal (CIP), caudate–CIP, and thalamus–frontoparietal (FRP) connections, while decreased FC was primarily found between subcortical regions and primary sensorimotor networks, including the auditory (AUD), sensorimotor hand (SMH), sensorimotor mouth (SMM), and visual (VIS) networks, along with subcortical–ventral attention (VEA) connections. Cortical networks were ordered along the sensorimotor-to-association axis^27^. For each cortical network, number of significantly increased and decreased FCs were calculated at 4-year follow-up, and the larger of the two was projected onto the cortical surface (top, left hemisphere). For subcortical regions, the total number of significantly altered FC for was estimated for each region at 4-year follow-up and then mapped onto the subcortical regions (bottom). (**B**) 50 FCs were significantly associated with PSU versus single substance use (SSU) at the 4-year follow-up, exhibiting similar but attenuated FC alterations compared with PSU versus NSU. (**C**) Only one FC was significantly associated with SSU versus NSU across time points. Statistical significance is indicated by asterisks (*P_FDR_ < .05; ** P_FDR_ < .01; *** P_FDR_ < .001, significant after false discovery rate (FDR) correction for 247 multiple comparisons). Heatmaps illustrated odds ratios (ORs) for associations from FC to PSU versus NSU, PSU versus SSU, and SSU versus NSU (blue = decreased connectivity; red = increased connectivity), and brain maps displayed the number of significant associations for each functional cortical network in the Gordon atlas and for subcortical regions derived from FreeSurfer automated segmentation (shown in purple). Abbreviations: rh, right hemisphere; lh, left hemisphere; VDC, ventral diencephalon; TH, thalamus; PT, putamen; PL, pallidum; HC, hippocampus; CD, caudate; CBC, cerebellum cortex; BS, brain stem; AG, amygdala; AB, accumbens area; CIO, cingulo-opercular; DEF, default; DOA, dorsal attention; NON, none; RET, retrosplenial temporal; SAL, salience; VIS, visual.

When comparing PSU with SSU, 50 connections were significantly associated at the 4-year follow-up (OR=0.79–1.24, P_FDR_<0.05; **Fig. 4b**), exhibiting similar but attenuated connectivity patterns relative to PSU versus NSU; whereas only one connection significantly differentiated SSU from NSU across time points (**Fig. 4c; Supplementary Table 19**).

Sensitivity analyses were conducted after excluding participants who had initiated PSU before each imaging time point (for example, for a participant who initiated PSU at year 5, only their baseline, year 2 and year 4 scans were included to predict subsequent PSU initiation). This analysis yielded similar effect sizes but reduced statistical significance (correlation R>0.89, P_FDR_<1.0×10^−10^ at all-time points), likely due to a reduced sample size (particularly at 4-year follow-up) (**Extended Data Fig. 6; Supplementary Table 20**). Associations between PRS_SUD_ and FC were largely not significant (**Extended Data Figs. 7–8; Supplementary Tables 21–22**). Additional analyses of cortical connectivity, and transition time in relation to lifetime substance use outcomes are presented in **Extended Data Figs. 9–10** and **Supplementary Tables 23–25**.

## DISCUSSION

To our knowledge, this is among the first large-scale, longitudinal population-based studies to integrate genetic liability, multidimensional environmental exposures and functional brain network development to examine factors associated with PSU initiation during adolescence. Using data from the ABCD cohort, we identified multilevel factors associated with PSU initiation and their transition time from NSU or SSU. We found that shared genetic liability across adult SUDs was associated with increased likelihood of PSU but not SSU initiation. Environmental exposures, including peer victimization, adverse life events, prenatal tobacco exposure, were associated with increased risk of PSU initiation, whereas parental monitoring, family cohesion and sleep duration were protective. These environmental associations were stronger among adolescents with higher genetic liability. In addition, adolescents with PSU showed differences in subcortical–cortical FC across development, particularly involving thalamic and caudate connections with frontoparietal and cingulo-parietal networks. Together, these findings suggest that adolescent PSU is associated with converging genetic, environmental and neurodevelopmental factors that are detectable early in development.

While previous studies have largely examined genetic risk for individual substances, growing evidence from adult samples indicates that SUDs share a substantial component of genetic liability^11,12^. The present findings extend this literature developmentally by showing that greater shared genetic liability for adult SUDs is associated with increased risk of PSU initiation, but not SSU, during adolescence. One possible interpretation is that PSU initiation during adolescence represents an early behavioral manifestation of shared genetic liability for adult SUDs, with higher liability youth more likely to progress beyond single-substance experimentation toward broader polysubstance involvement as opportunities arise. Broader exposure across substances may then create a risk-amplification process, in which greater opportunities to experience reinforcing effects increase continued use, further substance involvement, and eventual risk for SUDs. Although PRS_SUD_ explained only a modest proportion of variance, its selective association with PSU, but not SSU, supports the view that broad polysubstance involvement may index a generalized addiction-liability pathway rather than substance-specific risk alone. Thus, in this context, PRS_SUD_ is most informative as an etiologic marker linking adolescent PSU initiation to the shared genetic architecture of adult SUDs, rather than as a stand-alone tool for individual-level prediction.

This study also extends previous work linking adolescent substance use to individual environmental exposures, such as prenatal substance exposure^28^ and parental monitoring^15^, by examining a broad range of environmental factors simultaneously and assessing risk and protective factors specifically for PSU. Among the seven domains examined, life events or lifestyle, and peer relationships showed the strongest associations with PSU compared with both NSU and SSU. Exposure to stressful or adverse life events during childhood has been repeatedly associated with earlier initiation of substance use and increased likelihood of engaging in multiple substances^29^. Peer relationships also represent one of the most powerful influences on adolescent behavior, as adolescents may be more likely to initiate and escalate substance use in peer contexts where substance use is normative, accessible, or socially reinforced^30^.

At the same time, protective environmental factors may buffer against PSU initiation. Parental monitoring, family cohesion, and adequate sleep duration have consistently been linked to lower rates of substance use and delayed initiation of substance involvement^31,32^. These factors may reduce opportunities for substance exposure and shape adolescents’ attitudes toward substance use. Because several factors identified here are potentially modifiable, the present findings point to prevention targets across family, school, and community contexts. Interventions aimed at strengthening parental monitoring, improving family relationships, promoting healthier sleep patterns, and reducing exposure to high-risk peer contexts may be particularly relevant for youth at elevated vulnerability for polysubstance involvement^33^.

Beyond these main environmental associations, this study also identifies gene x environment interactions in PSU initiation. Adolescents exposed to greater adversity, including traumatic events, showed stronger associations between shared genetic liability for SUDs and PSU initiation, suggesting that adverse environments may amplify inherited vulnerability. Extending the risk-amplification framework described above, these findings are consistent with an adversity-amplified liability pathway, in which adolescents with elevated genetic risk may be more likely to use substances to regulate negative effects or trauma-related symptoms. In this context, polysubstance involvement may increase opportunities to identify substances that transiently alleviate distress, thereby reinforcing continued use through negative reinforcement mechanisms and potentially increasing risk for later SUDs^34^. These findings highlight that genetic liability does not act in isolation, but interacts with environmental context to shape adolescent substance-use trajectories.

This study also provides evidence that PSU initiation is associated with alterations in subcortical–cortical FC that become more pronounced across adolescence. Adolescents with PSU exhibited stronger connectivity between subcortical regions and association cortex, but weaker connectivity between subcortical and sensorimotor networks, relative to those with NSU. This pattern is broadly consistent with the sensorimotor-to-association (S-A) axis of cortical maturation^27^. During typical development, brain networks shift from subcortically anchored and sensorimotor-dominated organization toward greater integration within and between association cortices, which mature later and remain highly plastic during adolescence. Stronger subcortico–association coupling in adolescents with PSU may therefore reflect variation in the timing or trajectory of this developmental transition, potentially indicating divergent engagement of association-hub circuitry during sensitive developmental period. This interpretation is consistent with prior work implicating disrupted thalamo–cortical circuits in reward processing and addictive behaviors^35^. Viewed alongside the genetic and environmental findings, these functional-connectivity results suggest that PSU initiation is unlikely to reflect a single risk pathway. Rather, adolescent PSU may emerge from the intersection of shared genetic liability, social-contextual adversity and protection, and divergent maturation of subcortical–cortical circuitry relevant to reward, regulation, and executive control.

Several limitations should be considered when interpreting our findings. First, PSU in this study reflected initiation of more than one substance in the prior year (including single instances of use) and did not require frequent, heavy, or disordered use. Many adolescents who experiment with more than one substance do not progress to clinically significant SUDs or experience clinically meaningful impairment^36^. Accordingly, PSU should not be interpreted as a pathological phenotype, and language describing transitions from NSU or SSU to PSU is not intended to imply clinical escalation but rather to characterize heterogeneity in patterns of early substance experimentation. Second, substance-use outcomes were based primarily on self-report, which may introduce bias, because adolescents may underreport substance use behaviors. To address this concern, we also examined biological measures, including hair toxicology, which showed >78% concordance with self-reported substance-use outcomes across time points (**Supplementary Table 26**). Third, despite statistically significant associations, the variance in PSU explained by PGS was modest. These findings should therefore be interpreted as evidence of etiologic association rather than individual-level prediction. As GWAS sample sizes increase and become more diverse, future analyses may identify additional genetic associations with adolescent PSU and improve the portability of genetic findings across populations. Lastly, although this study leveraged longitudinal data, the observational design precludes causal inference, and unmeasured confounding may contribute to the observed associations.

In summary, this study provides converging evidence that genetic liability, environmental exposures, and brain network development are associated with adolescent PSU initiation. These multilevel factors differentiated adolescents who initiated PSU from those who initiated SSU, with differences in subcortical–cortical connectivity becoming more pronounced across adolescence. Together, these findings support an integrated developmental framework for understanding PSU initiation as heterogeneity in adolescent substance use involvement rather than as a pathological endpoint. Such a framework may help inform prevention strategies while maintaining a clear distinction between normative experimentation and clinically significant substance use.

## METHODS

### Study Design and Participants

This study used data from the ABCD Study (release 6.0), which enrolled 11,868 participants aged 9 to 10 years at baseline across 21 U.S. sites^22^. Release 6.0 includes the first tranche of year-6 follow-up data, with the remaining year-6 data available in subsequent releases^22^. Analyses included participants with available substance use outcomes within 6-year follow-up after genetic quality control (**Supplementary Methods**; **Extended Data Fig. 1**). Detailed cohort characteristics^22^ are summarized in **Table 1**. The ABCD Study received central institutional review board (IRB) approval from the University of California, San Diego, along with written parental consent and verbal assent from participants. This secondary analysis of de-identified ABCD data was exempt from IRB review at Mass General Brigham, Boston, U.S.

### Substance Use Outcomes

Substance use outcomes were ascertained from the Substance Use Interview, Substance Use Phone Interview (Mid-Year), and Hair Drug Toxicology (**Supplementary Table 27**). Participants were classified as cases for four binary lifetime substance use outcomes up to the 6-year follow-up (**Extended Data Fig. 2**): ALC (at least one full cup of alcohol consumed), NIC (any use of nicotine products, including cigarettes, e-cigarettes, smokeless tobacco, cigars, hookah, pipe, and nicotine replacements), CAN (use of marijuana products, including smoked or vaped flower, edibles, concentrates, and synthetic tetrahydrocannabinol products), and OTH (any use of other substances, including prescription drug misuse and illicit drugs). Three aggregated lifetime outcomes within the 6-year follow-up period were defined: NSU (reporting no lifetime substance use), SSU (lifetime use of only one type of substance at any follow-up time point), or PSU (concurrent use of at least two types over a one-year period)^1^, and these categories were mutually exclusive and collectively exhaustive. To leverage the longitudinal data, the transition time from NSU to PSU (or SSU) initiation was defined as the age at first PSU (or SSU) initiation minus baseline age, and from SSU to PSU initiation as the age at first PSU initiation minus the age at first SSU initiation (**Supplementary Table 2**).

### Psychopathology

Psychopathology was assessed annually by the parent/caregiver-reported Child Behavior Checklist (CBCL) total problems (raw scores),^37^ excluding the three items related to substance use (**Supplementary Methods**).

### Genotype Data, Quality Control, and Imputation

In the ABCD study, saliva and blood samples were collected at baseline for genotyping using the Affymetrix NIDA Smokescreen Array. Quality-controlled (QC) genotype and TOPMED-imputed data for 11,665 participants were downloaded from the NDA website. The procedures for genotyping, imputation, and QC in the ABCD study are detailed elsewhere^38^. Genetic principal components (PCs) and genetic relatedness were estimated, and participants were classified into five genetic ancestry groups—AFR (African/Admixed African American), AMR (Hispanic/Admixed Latin American), EUR (European), SAS (South Asian), and EAS (East Asian)—through clustering with 1000 Genomes Reference samples (**Extended Data Fig. 11**). Due to small sample sizes and few substance use cases in the SAS and EAS groups, subsequent analyses focused on the AFR, AMR, and EUR groups. For PGS analyses, we included imputed SNPs with imputation quality scores ≥ 0.8, minor allele frequency ≥ 1%, and Hardy-Weinberg Equilibrium *p*-value > 1×10^−6^.

### Genomic Structural Equation Modeling of General Substance Use

Building on recent multivariate GWAS studies^11^, we used GenomicSEM^39^ to construct a general addiction risk factor capturing common genetic vulnerability shared across five SUDs and improving polygenic prediction compared to single-trait GWAS^40^ (**Supplementary Table 6**): alcohol use disorder (AUD; sample N = 313,963, effective sample size [N_eff_]= 188,039)^41^, cannabis use disorder (CUD; N = 374,287, N_eff_ = 65,159)^42^, cocaine use disorder (COD; N = 8,463, N_eff_ = 6,285) ^43^, opioid use disorder (OUD; N = 447,950, N_eff_ = 179,212)^44,45^, and nicotine dependence (NIC; N = 244,890, N_eff_ = 39,420)^46^. Consistent with significant genetic correlations among these disorders (**Extended Data Fig. 12**; **Supplementary Table 28**), the common factor model fit the data well (χ^2^ = 4.10, p-value = 0.54, comparative fit index = 1, standardized root mean square residual = 0.03). The latent factor loaded significantly on all SUDs (standardized loadings on AUD = 0.84, CUD = 0.80, COD = 0.94, OUD = 0.98, NIC = 0.62, **Supplementary Table 7**).

### Generation of Polygenic Risk Scores (PRS)

We used PRS-CS^47^ to estimate posterior SNP effect weights. Parameters were set to a gamma-gamma prior (a = 1, b = 0.5), global shrinkage (phi) = auto, 1,000 MCMC iterations (500 burn-in), and thinning factor = 5. Ancestry-matched reference panels were derived from the 1000 Genomes Project. Using the estimated posterior SNP weights, we calculated PRSs for substance use disorders (PRS_SUD_) based on the latent addiction risk factor as described above. Consistent with GenomicSEM analysis, PRSs for individual SUDs were highly intercorrelated with PRS_SUD_ (**Extended Data Fig. 12**; **Supplementary Table 28**). We regressed out PRS_SUD_ from each disorder-specific PRS to index genetic risk specific to each SUD (PRS_AUD_, PRS_CUD_, PRS_COD_, PRS_OUD_, PRS_NIC_). PRSs were adjusted for population stratification using the top 10 genetic principal components within each ancestry group.

### Environmental Exposures

Following prior studies on environmental exposures associated with substance use^14,15,48,49^, and after retaining exposures with a prevalence ≥ 1% and variance inflation factor (VIF < 5) to mitigate multicollinearity (**Supplemental Methods**; **Extended Data Fig. 5**)^50^, 88 environmental exposures from the initial assessment were included, categorized into seven domains (**Supplemental Tables 29-30**): perinatal/early development (N = 11), prenatal substance exposure (N = 5), life event/lifestyle (N = 22), family (N = 29), neighborhood (N = 8), school (N = 3), and peer relationship (N = 10).

### Functional Connectivity

Baseline, 2-year, 4-year, and 6-year follow-up resting-state functional magnetic resonance imaging (fMRI) were acquired and preprocessed (**Supplemental Methods**)^26^. Post-processed data were used to characterize functional connectome, including Fisher Z-transformed averaged correlations within and between 13 Gordon cortical networks, and between 13 cortical networks and 19 subcortical regions^26^. For quality control, only scans that passed quality control provided by the ABCD Study^26^ and had mean framewise displacement (FD)<0.2 mm were included (**Extended Data Fig. 1**).

### Statistical Analyses

To examine whether PSU identifies youth at ultra-high risk for psychopathology, linear mixed-effects models (LMMs) were used to examine associations from lifetime substance use to the CBCL total problems at each time point. Age, sex, pubertal stage, income-to-needs ratio (INR, a global measure of socioeconomic status), and genetic ancestry were included as fixed-effect covariates, and family ID and enrollment site were modeled as random effects to account for within-individual, familial, and site-level variability. LMMs were also used to examine associations from PRS_SUD_ to FC, adjusting for the same covariates as well as mean FD to account for image quality, and using the same random effects along with scanner manufacturer to account for inter-scanner variability.

Generalized linear mixed-effects models (GLMMs) with binomial error structure and logit link function were used to examine associations between lifetime substance use and (a) six PRSs, (b) 88 environmental exposures, and (c) FC measures at each time point, adjusting for the same covariates and random effects as previously described. The PRS analysis was additionally stratified by genetic ancestry group, while the environmental and FC analyses adjusted for genetic ancestry as a covariate. Additional ancestry-stratified models were fit to assess PRS_SUD_-by-environment interactions on lifetime substance use, by defining a binary PRS_SUD_ variable that classified participants into a high-genetic-risk group (top 25th percentile of PRS_SUD_) and low-genetic-risk group (bottom 75th percentile of PRS_SUD_) levels within each ancestry group following previous studies^24,25^, also adding PRS × covariate and environment × covariate interactions to reduce spurious genome-wide signals^51^.

Finally, Cox proportional hazards models were used to examine associations between PRS, environment, and baseline FC and time to substance use initiation. Models were fit separately for time to transition from NSU to PSU (or SSU) initiation, and time from SSU to PSU initiation, adjusting for age, sex, pubertal stage, and INR, and random effects for family and site. As before, the PRS model was stratified by ancestry group, while environment analyses additionally adjusted for ancestry group, and FC analyses further adjusted for ancestry group, mean FD, and scanner.

For all analyses, covariates were aligned with the assessment time of independent variables, and numeric variables were standardized. Standardized coefficients (β) and associated two-tailed p-values from LMMs for continuous outcomes, adjusted odds ratios (ORs) with 95% confidence intervals (CIs) from GLMMs for binary variables, and hazards ratios (HRs) with 95% CIs from Cox proportional hazards models were reported. False discovery rate (FDR) of 0.05 was used to declare significant findings for multiple comparisons across 7 time points for CBCL total problems, 3 × 6 PRS associations (ancestry-by-PRS), 88 environmental exposures, and 247 subcortical-cortical connections, respectively. Analyses were conducted in R (version 4.4.0) with the *lmerTest*, *glmmTMB*, and *coxme* packages.

## Acknowledgements

PL was supported by grants from the U.S. National Institute of Health (R01MH119243, R01GM148494) and by MassGeneral Brigham Department of Psychiatry. JMG is supported by K02DA052684 and R01DA051540. SEH is supported by R01DA057566, K18DA059913, and R34DA062280. JLR is supported by R01MH124694. The ABCD Study is supported by the National Institutes of Health (NIH) and additional federal partners under award numbers U01DA041048, U01DA050989, U01DA051016, U01DA041022, U01DA051018, U01DA051037, U01DA050987, U01DA041174, U01DA041106, U01DA041117, U01DA041028, U01DA041134, U01DA050988, U01DA051039, U01DA041156, U01DA041025, U01DA041120, U01DA051038, U01DA041148, U01DA041093, U01DA041089, U24DA041123, U24DA041147. Additional support for this work was made possible from NIEHS R01-ES032295 and R01-ES031074. Funding supporting this study was provided by the Spanish ‘Ministerio de Ciencia, Innovación y Universidades’ (PID2021-1277760B-I100 and PID2024-158634OB-I00, to BC and NFC), ‘Generalitat de Catalunya/AGAUR’, (2021-SGR-01093, to BC and NFC), ICREA Academia 2021 (to BC), ‘Fundació La Marató de TV3′ (202218-31, to BC) and ‘Ministerio de Sanidad, Servicios Sociales e Igualdad/Plan Nacional Sobre Drogas’ (PNSD-2020I042 and PNSD-2024|056, to NFC). A full list of supporters is available online. A list of participating sites and a complete list of the study investigators can be found online. ABCD consortium investigators designed and implemented the study and provided data but did not necessarily participate in analysis or writing of this report. This manuscript reflects the views of the authors and does not reflect the opinions or views of the NIH or ABCD consortium investigators. The ABCD data repository grows and changes over time. The ABCD data used in this report came from https://doi.org/10.82525/jy7n-g441. We would like to thank the research participants and employees of 23andMe, Inc. for making this work possible.

## Author Contributions

PHL, DZ, and BS have directly accessed and verified the ABCD data reported in the manuscript. PHL and JG conceptualized the idea and designed the study. PHL, DZ, JG, DJ, and BS were involved in data curation, formal analysis, investigation, visualization, and validation of the findings. PHL and JG were involved in funding acquisition, supervision, and project administration. PHL, JG, DZ, and BS wrote the original draft. All authors were involved in the critical review and editing. All authors approved the final version of the manuscript.

## Declaration of Interests

All other authors have no interest to declare.

## Role of the Funder/Sponsor

The funding sources had no role in the design and conduct of the study; collection, management, analysis, and interpretation of the data; preparation, review, or approval of the manuscript; and decision to submit the manuscript for publication.

## Data Sharing

The Adolescent Brain Cognitive Development data used in this report are available from the NIH Brain Development Cohorts (NBDC) Data Hub (DOI: https://doi.org/10.82525/jy7n-g441) to Authorized Users. We used open-source software packages from R, and publicly available methods, GenomicSEM, PLINK, and PRS-CS (**Supplementary Table 6**).

## Notes

### Competing Interest Statement

The authors have declared no competing interest.

### Funding Statement

PL was supported by grants from the U.S. National Institute of Health (R01MH119243, R01GM148494) and by Mass General Brigham Department of Psychiatry. JMG is supported by K02DA052684 and R01DA051540. JLR is supported by R01MH124694. The ABCD study is supported by the National Institutes of Health (NIH) and additional federal partners under award numbers U01DA041048, U01DA050989, U01DA051016, U01DA041022, U01DA051018, U01DA051037, U01DA050987, U01DA041174, U01DA041106, U01DA041117, U01DA041028, U01DA041134, U01DA050988, U01DA051039, U01DA041156, U01DA041025, U01DA041120, U01DA051038, U01DA041148, U01DA041093, U01DA041089, U24DA041123, U24DA041147. Additional support for this work was made possible from NIEHS R01-ES032295 and R01-ES031074. Funding supporting this study was provided by the Spanish Ministry of Science, Innovation, and Universities (PID2021-1277760B-I100 and PID2024-158634OB-I00, to BC and NFC), Government of Catalonia/AGAUR, (2021-SGR-01093, to BC and NFC), ICREA Academia 2021 (to BC), TV3 Marathon Foundation (202218-31, to BC) and Ministry of Health, Social Services, and Equality/National Plan on Drugs (PNSD-2020I042 and PNSD-2024|056, to NFC).

### Author Declarations

This study utilizes publicly available data from the Adolescent Brain Cognitive Development (ABCD) Study. The ABCD Study is a nationwide clinical research initiative involving thousands of adolescents across 21 sites in the United States. Since the data used in this project are de-identified and publicly accessible, the research conducted using ABCD data qualifies for IRB exemption.

### Summary of Updates

We have revised the manuscript in response to the reviewers' comments by adding new analyses, clarifying the study design and interpretation of the findings, and further refining the presentation of the results and discussion.

